# Prenatal carrier screening for spinal muscular atrophy among Thai pregnant women

**DOI:** 10.1101/2024.04.17.24305978

**Authors:** Chayada Tangshewinsirikul, Panyu Panburana, Maneerat Prakobpanich, Takol Chareonsirisuthigul, Donniphat Dejsuphong, Thipwimol Tim-aroon, Chaiyos Khongkhatithum, Thanyachai Sura, Atchara Tunteeratum, Duangrurdee Wattanasirichaigoon

## Abstract

**Objectives:** To investigate the acceptance rate for spinal muscular atrophy (SMA) carrier screening among Thai pregnant women, their attitudes toward the prenatal screening, carrier rate, and the frequencies of *SMN2* copy numbers.

**Materials and methods:** Singleton pregnant women who aged ≥18 years, with a gestational age of ≤14 weeks at their first visit, were invited to participate the study. All participants completed the questionnaire: Section I - demographic data. Then, they received a pre-test group counseling, followed by an offer of SMA carrier testing at no cost and completion of the questionnaire: Section II - awareness and attitudes toward the screening and Section III – reasons for their choosing ‘to have’ or ‘not to have’ the screening done. Only those having the test done and undergoing post-test counseling were asked to complete the questionnaire: Section IV – attitudes toward the screening process.

**Results:** We found a high acceptance rate for carrier screening at 91.4% (181/198 participants), a carrier rate of 2.2% (1 in 45), and high frequency of ≤2 copies of *SMN2* (98.3%). The preexisting knowledge about SMA was low (30.8%). The majority of participants became realized about the severity of SMA and its burden to the families (94.4%) and agreed to have fetal diagnosis if they were found to be a couple-at-risk (92.4%). Most participants (98%) suggested that SMA carrier screening should be offered to all pregnant women and that the cost of testing should be covered by the government and/or by their health coverage schemes (95.5%).

**Conclusion:** The high acceptance rate and positive attitude toward prenatal SMA carrier was demonstrated among Thai pregnant women. Data from the present study highlight urgent needs for endorsement from professional society and public health policy in advancing the SMA carrier screening program in Thailand.

## Introduction

Spinal muscular atrophy (SMA) is a severe neuromuscular disorder which results in progressive muscle weakness, with an estimated incidence of 1 in 6,000-10,000 births worldwide [1]. SMA is divided into four clinical types depending on the age of onset and clinical progression. Type I SMA (Werdnig–Hoffmann) has clinical onset within the first three months of life, usually leading to respiratory failure and death by two years of age. The onset of type II SMA is 6-18 months of age, and the affected children may sit independently but cannot stand or walk, with 70% surviving beyond 25 years of age. Patients with type III SMA (Kugelberg– Welander), with the onset at ≥18 months, have difficulty climbing stairs and running and often have a normal lifespan. Type IV SMA or adult-onset SMA is described with slow progression of muscle weakness beginning at around 30 years of age, and with no reduction in life expectancy [2]. Type I SMA has the highest incidence among SMA, at 50-60% [2-5]. However, there is a marked difference between incidence and prevalence of type I SMA because of its low survival [6-7]. Among the living individuals with SMA, types II and III together account for 75-83% [6-9].

Survival motor neuron 1 (*SMN1*) gene, located on 5q11.2–q13.3, is the disease-causing gene of SMA. Approximately 95-98% of SMA cases are caused by a homozygous deletion of exon 7 or exons 7-8 of the *SMN1* gene, with 3%–5% being due to point mutation or compound heterozygosity between the deletion and point mutations [2]. *SMN2* is a highly homologous gene of *SMN1*, they are located next to each other. The complete absence of *SMN2* yields no clinical effect on healthy individuals. The copy number of *SMN2* rather modifies the severity of SMA [2, 6, 10]. Most people have two copies of *SMN1*, each on the homologous chromosome, and two copies of *SMN2*, each on the homologous chromosome as well [2,11], The higher number of *SMN2* copy number is associated with less severity of the SMA phenotypes (1 copy of SMN2: 96% - type I SMA; 2 copies: 79% - type I; 3 copies: 54% - type II and 31% - type III/IV; ≥4 copies: 88% - type III/IV) [2]. Therefore, simultaneous copy number detection of *SMN1-*exon 7 (or exons 7-8) and *SMN2*-exon7 is widely used for molecular diagnosis and prognostication for SMA [2,12]. As for carrier screening in general population, copy number analysis of the *SMN1* is a preferred method, owing to its high throughput, rapid turn-around time, and economical reason [13-15].

Carrier frequency of SMA in general population is high at approximately 1 in 33-94 worldwide [2, 11, 13, 15, 16], and potentially with the most severe phenotype due to the high frequency of ≤2 copies of *SMN2* among the carriers [15, 17-21]. Screening for SMA carrier before conception or early in pregnancy, regardless of religion or ethnicity, has been recommended by the American College of Medical Genetics (2008) and by the American College of Obstetricians and Gynecology (2017) [22, 23].

Thailand is a middle-income country with well-established universal health coverage (UHC) since 2002 (https://eng.nhso.go.th/) Almost all Thai citizens are currently covered by one of the three health benefit schemes: 74% under the Thai UHC, 7% covered by the Civil Servant Medical Benefit Scheme (CSMBS, for government/state employees), and 19% under the Social Security Scheme (SSS for working adults). Notably, 93% of Thai Children are covered by the UHC. Prenatal screening for thalassemia (carrier rate 40%) has been offered as the national policy since 1992, with a good coverage at 98% of pregnant women [24, 25]. Carrier frequency of SMA among Thai population is 1 in 40 [26, 27], but SMA carrier screening is not included in the national program or routine antenatal care in the country. Newborn screening for SMA is not offered and specific treatments for SMA (such as mRNA-modifying treatment and gene therapy) is not reimbursable under the existing health schemes in the country. Carrier testing for SMA and reproductive choices are offered to only the families with previous affected child/relatives with SMA.

To explore the potentials for establishing carrier screening for SMA during prenatal period, we set off to conduct this study. Our objectives were to assess the acceptance rate of the carrier screening among Thai pregnant women, to evaluate their attitudes toward the screening, to determine the prevalence of SMA carrier and the frequencies of *SMN2* copy numbers.

## Materials and Methods

### Study design and participants

This was a prospective and descriptive study, conducted during 10 March 2021 and 31 March 2022, at the antenatal care clinic, Ramathibodi Hospital. The protocol was granted by the Ramathibodi Hospital Institutional Review Board (COA, MURA2020/1420) and complied with the Declaration of Helsinki. The study was registered as a clinical trial (registration number NCT04859179).

Eligible participants were women with singleton pregnancy, who were aged ≥18 years and capable of reading and understanding Thai, and with a gestational age of ≤14 weeks at their first visit with our antenatal care clinic. They were invited to enroll in the study, by one of the authors (MP) and asked to provide a written informed consent. Exclusion criteria were refusal to give a written consent, withdrawal during or after the completion of the study, and insufficient data obtained from the questionnaire.

### Questionnaire

A Thai-language questionnaire was developed using information from a literature review, then validated by six clinical experts: two maternal and fetal medicine specialists, two pediatric geneticists, and two clinical scientists. The reliability of the questionnaire was 0.797, as assessed by using Cronbach’s alpha.

The questionnaire contained four sections, I-IV. Section I focused on the participants’ demographic data, including age, education, occupation, religion, and gravida and previous history of fetal anomalies. Section II asked about awareness and attitudes toward SMA carrier screening. This section comprised of nine question items, including the awareness of severity of SMA and its burden on the patients and families, the usefulness and interest in having prenatal carrier screening for SMA and their confidence in the reliability of the test offered, agreement in having definitive test for fetal diagnosis, possible considering termination of pregnancy as an alternative if the fetus to be found affected, opinion on having SMA carrier screening as part of a standard antenatal care, and their willingness to self-pay for the screening test. The answer options were ‘agree’, ‘not sure’, and ‘disagree’.

As for Section III, we explored the reason(s) and/or rationale thinking of the participants for choosing ‘to have’ or ‘not to have’ the carrier screening test. Multiple choice answers were provided to choose and the participants could choose more than one answer as wanted. We also asked the participants’ opinions on a reasonable price for the screening test if it had to be self-pay, and who (individuals, organizations, health schemes, or else), in their opinion, should be responsible for cost of testing if the screening would become part of standard antenatal care in the country.

Section IV consisting five question items, was about the participants’ attitudes and reflections of their immediate past experience with going through the SMA carrier screening process including pre-test and post-test counselling: overall feeling/satisfaction to the screening process, the usefulness of the information provided, the essence of post-test counseling, the whole process is time worthy, and that the carrier screening should be offered to all pregnant women during antenatal visit. The answer options for each question were ‘agree’, ‘not sure’, and ‘disagree’.

### Data collection

All the participants, after the completion of Section I of the questionnaire, were provided with pre-test group counseling delivered by trained maternal and fetal medicine specialists using a PowerPoint presentation which lasted for about 10 minutes. The participants then received the Thai-language brochure to read in a designated area, this was to allow the participants time to digest the information received and self-check of their understanding. The slide presentation for the group counseling and the brochure contained similar information, including the incidence, clinical presentations, natural course and outcomes of SMA; treatment options and the efficacy of available treatments, affordability of the treatment in the country; carrier rate and carrier screening; and prenatal diagnosis and its downstream services.

After pre-test counseling, a free-of-charge SMA carrier testing (as part of this research) was offered to all participants and they were asked to complete the questionnaire Sections II and III, regardless of their choosing to have the test done or not. A sequential prenatal SMA carrier screening protocol was established to use in this research (Figure 1). For those choosing not to have the test done, the study ended at this point. About those choosing to have carrier testing performed, 10-ml of venous blood sample was taken for *SMN1* and *SMN2* analysis.

**Figure 1.**
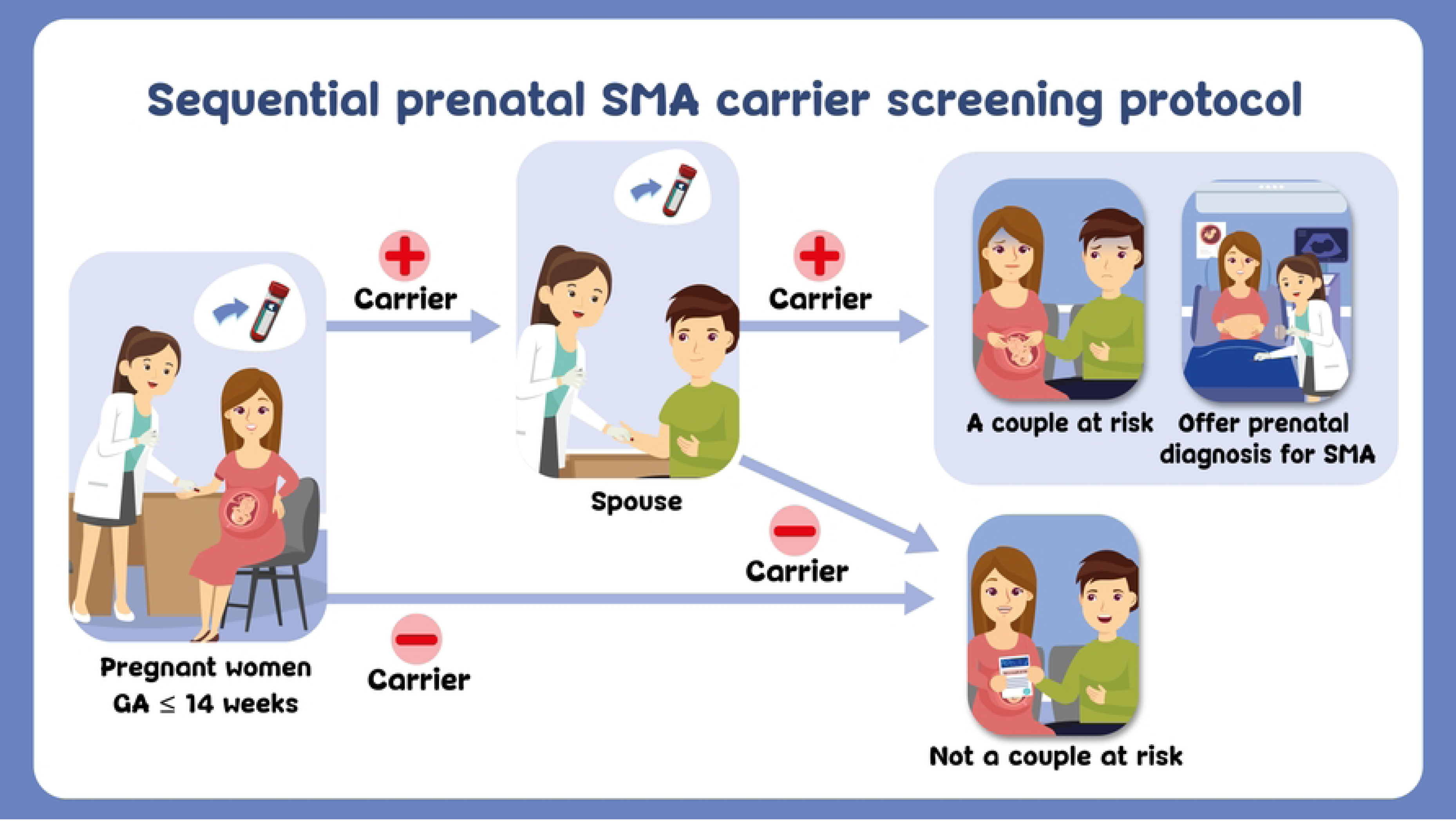
Sequential prenatal spinal muscular atrophy (SMA) carrier screening protocol.

Post-test counseling for each individual was offered by a maternal and fetal medicine specialist at an antenatal care clinic, upon the result became available. Then, the participants were requested to finalize the Section IV of the questionnaire. For those who were found to be a carrier, their partner was counseled and offered the carrier testing (with no charge). Prenatal diagnosis would be discussed to a couple at risk.

### Carrier testing and copy numbers analysis for SMN1 and SMN2

To detect copy number of *SMN1* and *SMN2*, we performed multiple ligation-dependent probe amplification (MLPA) analysis, using a commercially available MLPA kit for SMA (SALSA P021), following the manufacturer’s protocol (MRC Holland, Amsterdam, the Netherlands). The MLPA reactions included internal quality controls and negative controls. The PCR products were analyzed using the Applied Biosystems 3500 Genetic Analyzer (Thermofisher Scientific, MA, USA) and Coffalyser.net software (MRC Holland), following the manufacturer’s instructions. The tests were performed at the Human Genetics Laboratory, Ramathibodi Hospital. Carrier status, in the present study, is defined as having only 1 copy of *SMN1* exon 7. The results of carrier testing were reported to the clinician researchers (CT, PP, and MP) within two weeks.

### Statistical analyses

Categorical variables were described as the number (percent). Parametric continuous variables were expressed as a mean and standard deviation.

## Results

A total of 237 pregnant women were assessed, 35 did not meet the inclusion criteria and four declined to participate, leaving 198 (83.5%) of eligible participants to be enrolled in the study. Among 181 out of 198 (91.4%) research participants who took the screening test, four were identified as carriers of SMA. Their partners were screened, and none was found to be a carrier, resulting in no couple at risk detected in the present study (Figure 2).

**Figure 2.**
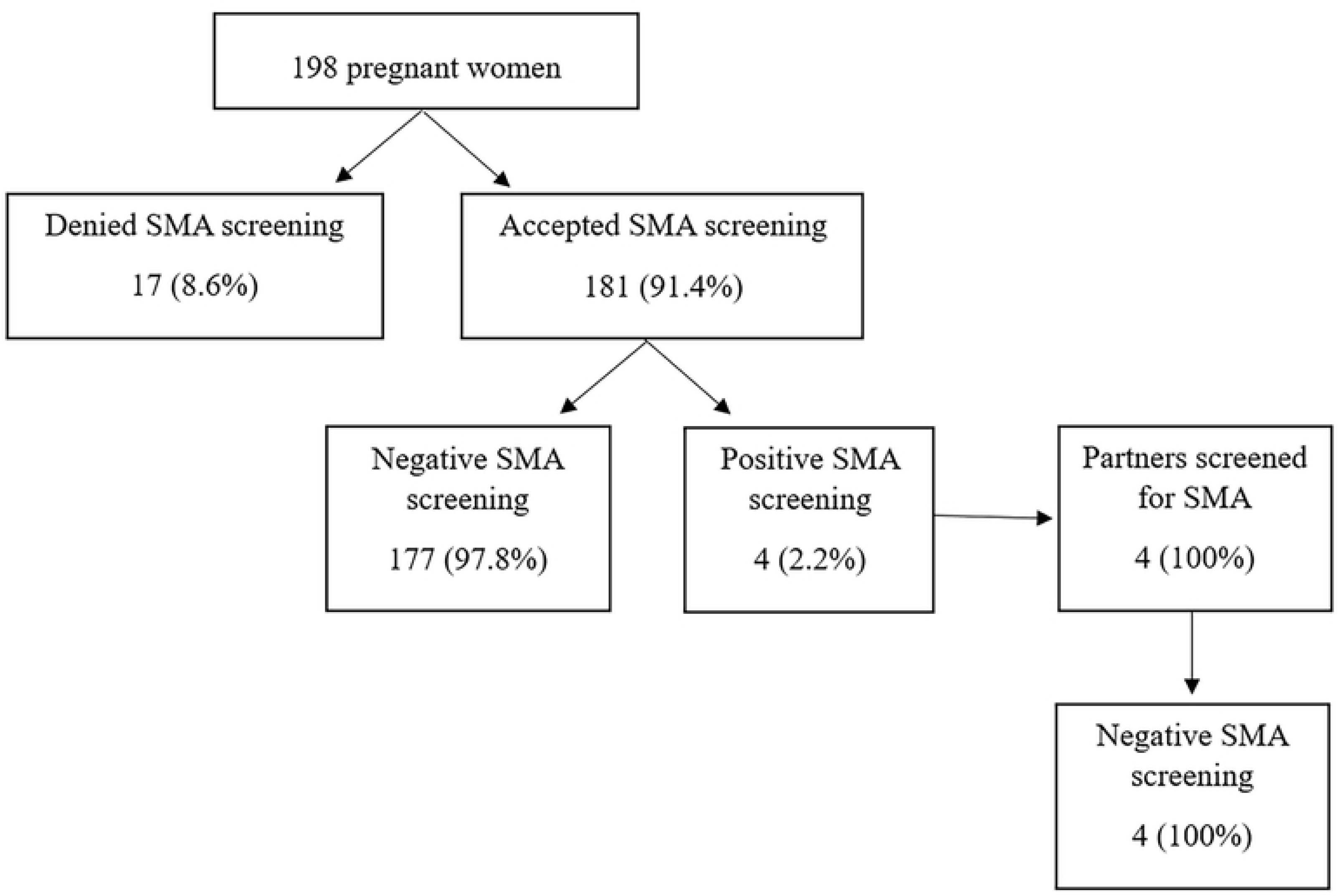
Prenatal carrier screening among 198 pregnant women.

The mean age of the participants was 32 ± 5.1 years and over half (66.2%) of them had a Bachelor’s degree or higher (Table 1). None had a child or family relative(s) with SMA. Two-thirds of the participants said that they had never heard of SMA before.

### Acceptance and attitudes toward SMA carrier screening

Around 95% or over of the participants agreed that SMA is a serious disorder resulting in severe life suffering, and that the carrier screening could reduce unnecessary anxiety during pregnancy and it should be offered to all pregnant women (Table 2). About 80-92% or more of the participants expressed that having a child with SMA would complicate their life, they were not reluctant to have the screening test done, they had a good faith in the accuracy of the test, and they would have fetal diagnosis if they would be found to be a couple at risk (Table 2). Almost half of the participants expressed their tendency to choose termination of pregnancy for an affected fetus (Table 2).

In the present study, the most common reasons for having the carrier screening included wanting to know their carrier status (87.8%), being worried to have an affected fetus (72.9%), and no extra cost for the test (50.3%) (Table 3). As for those declining the test, they reasoned that SMA was rarely found (58.8%) (Table 3).

Almost half of the participants (47.7%) said that they could afford 501–1,000 Baht (∼14-28 USD) if self-pay for the SMA carrier screening. Most participants (95.5%) thought that the cost of carrier testing should be covered by the government and/or their health coverage schemes (Table 4).

After receiving the result of screening test, all the participants expressed the feeling of glad that they had had the test done. They all agreed that the information about SMA provided to them were very useful and that post-test counseling was essential (Table 5).

### SMA carrier rate and frequencies of SMN2 copy number

Only 2.2% (4/181) of the participants were found to be carrier (Table 6). As for the *SMN2*, the majority of participants had 2 copies (58.5%), followed by ≤1 copy (39.8%), and ≥3 copies (1.7%). Regarding the carrier individuals, three had 2 copies and the other had 1 copy of *SMN2*.

## Discussion

We found a high acceptance rate for prenatal SMA carrier testing (91.4%) and positive attitudes towards the screening test, among the pregnant women in this study. Most participants (92%) would choose fetal diagnosis if they were found to be a couple at risk and consider having interruption of pregnancy in case of affected fetus (47%), these finding were in agreement with earlier studies [13, 15]. The majority of the participants suggested SMA carrier testing to be part of standard antenatal care and covered by the public health system.

The low awareness of SMA among the participants, were probably due to the rarity of the disorder; being a hidden problem of SMA owing to the short life of significant proportion of the patients, the patients’ low social activities because of respiratory and ambulatory difficulties; and the lack of public health education and screening services for SMA [9, 28, 29].

Despite of generally low awareness of SMA prior to the study, the acceptance rate for SMA prenatal carrier screening was high among the participants in the present study, likely due to several factors including the knowledge about SMA given, perceived usefulness of the pre-and post-test counseling provided, trust in the test offered, testing at no extra cost (Table 3), and availability of the downstream services. Some studies have shown a significantly increased acceptance rate if covered by medical insurance/health scheme (97.8%) compared to self-funded screening (81.1%) [29, 30]. Other influencing factors included race, parity, and religion [17, 29].

As for those declining the screening test, the main reasons were the low prevalence of SMA (58.8%) and not choosing interruption of the pregnancy for an affected fetus (17.6%). Our findings were consistent with previous reports that people without SMA experience had significantly lower rate of taking carrier testing because they had a dimmer view of the condition than those with experiential knowledge [29, 31]. Therefore, the endorsement from public health authority and professional society and effective mass communications are urgently needed in advancing the SMA carrier screening program [29-31].

The SMA carrier rate found in this study was 1 in 45, with estimated prevalence of SMA at 1 in 8,264, similar to those found worldwide [11, 13, 15, 16, 17-21, 26, 27]. The majority (98.3%) of the participants had ≤2 copies of *SMN2*, suggesting that a major proportion of Thai affected individuals (fetuses) would be predicted to have the most severe phenotype (type I) of SMA, consistent with other studies [2, 11].

To establish a new prenatal carrier screening program in any country, it is essential to have testing that is acceptable to the target population, with timely turnaround time, appropriate pre-test and post-test counseling, and affordable downstream services including testing for the partners, fetal diagnosis, and choices of continuation and interruption of pregnancy. Other local context, such as affordable treatment and predicted severity of the disorder, would also tremendously influence the individual’s decision on taking or declining the testing. Therefore, population-specific data, both biomedical (carrier rate, copy number of *SMN2*, type of SMA, availability of specific treatment) and sociocultural aspect (religion and extended socio-educational supports) are highly essential for policy planning for each country.

SMA fits all the criteria for prenatal carrier screening, as follows: 1) severity of the disorder; 2) high carrier frequency in the general population more than 1 in 100; 3) available carrier screening test with a high sensitivity and specificity; 4) availability of prenatal diagnosis during early gestation; and 5) timely turnaround time for the screening test and fetal diagnosis [22, 28].

The impact of population-based carrier screening for SMA has been well shown in Israel by that 31% of the cases were prenatally diagnosed following the introduction of the screening through healthcare service and insurance plans (since 2008), and even higher at 52% after it became the national program (since 2013) [30].

To date, Thailand has had a successful prenatal carrier screening program for thalassemia using a sequential testing strategy. Both thalassemia and SMA are inherited in an autosomal recessive fashion. We believe that it is highly pragmatic to incorporate the SMA screening along with thalassemia screening program for Thai population. However, public education of SMA and additional training for healthcare professionals especially those in family medicine and prenatal setting are required. Moreover, the SMA carrier screening can be further expanded to the pre-conceptional period.

To our knowledge, this study is the first study to evaluate the acceptance of prenatal SMA carrier screening and its influencing factors among Thai population, using a prepared set of educational materials and counseling program. The study is one of a very few studies of its kind, among Asian populations and low-middle income countries which is underrepresented in the literature. The validated Thai-language brochure and the whole process of testing strategy which were approved for its usefulness by the participants could be applied to a large-scale screening among Thai population. Lastly, this is the first demonstration of copy number of *SMN2* among general population of Thais.

The authors appreciated limitations of the study, including a small sample size, the absence of couples at risk and affected fetus identified, which might have resulted in different answers on choosing fetal diagnosis and pregnancy outcome.

## Conclusion

The acceptance rate for prenatal SMA carrier screening was high among Thai pregnant women and it could be further amplified by effective mass education to general population and affordable screening services. The estimated prevalence and carrier frequency of SMA among Thai population was found equivalent to the worldwide data and the majority of patients were predicted to have the most severe type of SMA, necessitating the needs for national program for disease prevention. Population-specific data, both biomedical and sociocultural aspects, are markedly important for national policy planning for the carrier screening.

## Acknowledgements

We are thankful of the Faculty of Medicine Ramathibodi Hospital, Mahidol University for providing Research Career Development Awards to DW, TT, and DD. We thank Melissa Leffler, MBA, from Edanz (www.edanz.com/ac) for editing a draft of this manuscript.

## Authors’ contributions

Conceptualization: Chayada Tangshewinsirikul, Takol Chareonsirisuthigul, Donniphat Dejsuphong, Duangrurdee Wattanasirichaigoon

Data curation: Chayada Tangshewinsirikul, Maneerat Prakobpanich, Takol Chareonsirisuthigul

Formal analysis: Chayada Tangshewinsirikul, Takol Chareonsirisuthigul, Duangrurdee Wattanasirichaigoon

Funding Acquisition: Chayada Tangshewinsirikul

Investigation: Chayada Tangshewinsirikul, Panyu Panburana, Maneerat Prakobpanich, Takol Chareonsirisuthigul, Donniphat Dejsuphong, Thipwimol Tim-aroon, Duangrurdee Wattanasirichaigoon

Methodology: Chayada Tangshewinsirikul, Panyu Panburana, Maneerat Prakobpanich, Takol Chareonsirisuthigul, Donniphat Dejsuphong, Thipwimol Tim-aroon, Chaiyos Khongkhatithum, Thanyachai Sura, Atchara Tunteeratum, Duangrurdee Wattanasirichaigoon

Supervision: Duangrurdee Wattanasirichaigoon

Writing– original draft: Chayada Tangshewinsirikul, Takol Chareonsirisuthigul

Writing– review & editing: Duangrurdee Wattanasirichaigoon

## Data availability statement

The datasets generated and/or analyzed during the current study are not publicly available due to limitations of ethical approval involving the patient data and anonymity but are available from the corresponding author on reasonable request.

